# Genetic constraint at single amino acid resolution improves missense variant prioritisation and gene discovery

**DOI:** 10.1101/2022.02.16.22271023

**Authors:** Xiaolei Zhang, Pantazis I. Theotokis, Nicholas Li, the SHaRe Investigators, Caroline F. Wright, Kaitlin E. Samocha, Nicola Whiffin, James S. Ware

**Affiliations:** National Heart & Lung Institute, Imperial College London, London W12 0NN, United Kingdom; MRC London Institute of Medical Sciences, Imperial College London, London W12 0NN, United Kingdom; Royal Brompton & Harefield Hospitals, Guy’s and St. Thomas’ NHS Foundation Trust, London SW3 6NP, United Kingdom; University of Exeter Medical School, Institute of Biomedical and Clinical Science, Royal Devon and Exeter Hospital, Exeter EX2 5DW, UK; Center for Genomic Medicine, Massachusetts General Hospital, Boston, MA 02114, USA; Program in Medical and Population Genetics, Broad Institute of MIT and Harvard, Cambridge, MA 02142, USA; Wellcome Centre for Human Genetics, University of Oxford, Oxford OX3 7BN, UK; Division of Cardiovascular Medicine, Stanford University Medical Center, Stanford, CA, USA; Department of Cardiology, Boston Children’s Hospital, Boston, MA, USA; Division of Cardiovascular Medicine and Penn Cardiovascular Institute, Perelman School of Medicine, University of Pennsylvania, Philadelphia, USA; Department of Internal Medicine-Cardiology, University of Michigan, Ann Arbor, MI, USA; Cardiovascular Division, Brigham and Women’s Hospital, Boston, MA, USA; Centre for Population Genomics, Garvan Institute of Medical Research, and UNSW Sydney, Sydney, Australia; Centenary Institute, The University of Sydney, Sydney, Australia; Department of Cardiology, Royal Prince Alfred Hospital, Sydney, Australia; Department of Internal Medicine, Yale University, New Haven, CT, USA; Department of Cardiology, Thoraxcenter, Erasmus MC Rotterdam, Rotterdam, Netherlands; Cardiomyopathy Unit, Careggi University Hospital, Florence, Italy; Heart Institute (InCor), University of Sao Paulo Medical School, Sao Paulo, Brazil; Children’s Hospital of Philadelphia, PA, USA; Agnes Ginges Centre for Molecular Cardiology Centenary Institute, The University of Sydney, Australia; Sydney Medical School, Faculty of Medicine and Health, The University of Sydney, Australia; Cincinnati Children’s Hospital Medical Center, Heart Institute, Cincinnati, OH, USA

## Abstract

The clinical impact of most germline missense variants in humans remains unknown. Genetic constraint identifies genomic regions under negative selection, where variations likely have functional impacts, but the spatial resolution of existing constraint metrics is limited. Here we present the Homologous Missense Constraint (HMC) score, which measures genetic constraint at quasi single amino-acid resolution by aggregating signals across protein homologues. We identify one million possible missense variants under strong negative selection. HMC precisely distinguishes pathogenic variants from benign variants for both early-onset and adult-onset disorders. It outperforms existing constraint metrics and pathogenicity meta-predictors in prioritising *de novo* mutations from probands with developmental disorders (DD), and is orthogonal to these, adding power when used in combination. We demonstrate utility for gene discovery by identifying seven genes newly-significant associated with DD that could act through an altered-function mechanism. Overall, HMC is a novel and strong predictor to improve missense variant interpretation.

## Main

Quantifying the depletion of natural variation in human populations provides a powerful approach to identify variants of large effect^1–8^. Since variants causing severe early-onset disorders are under selective pressure, they are observed less often than functionally neutral variants. Such depletion of genetic variation (constraint) has been shown to provide strong evidence to prioritise disease-associated genes^1–3^, identify critical regions within genes^4,5^, and investigate the effect of non-coding variants^6–8^.

However, these existing metrics have limited resolution to analyse individual residues, and limited application in genes with sparsely distributed pathogenic missense variants since they explicitly rely on signals clustered linearly within genes^1–5^ (**Figure S1**). To address this issue, we sought to develop an amino-acid level constraint metric. Since we are still underpowered to evaluate the depletion of variants at individual residues, we evaluate homologous residues instead. While previous studies have aggregated variant information over homologous residues to infer functional effect^9–14^, a pathogenicity predictor using genetic constraint hasn’t been studied.

Here we developed an amino-acid level constraint metric by aggregating the signal over evolutionarily equivalent positions across human protein domains. While there are alternative definitions of homology, we use protein domain families defined by the Pfam database^15^, which identifies regions of homology in most genes (see **Supplementary Information** for discussion on alternative approaches). Of 70 million possible missense variants (defined by NCBI RefSeq Select transcripts^16^) in the human genome, 28 million are mapped to Pfam protein domain families. After excluding residues with limited statistical power (see **Method**), 15 million rare missense variants (∼21% of all possible missense variants) are assessable, which are defined as those with a minor allele frequency (MAF) < 0.1 or absent from the Genome Aggregation Database (gnomAD v2.1.1; 125,748 samples)^2^. Given a set of homologous residues, we calculated the genetic intolerance of missense variants at individual residues as the ratio of the number of rare missense variants observed in the 125K gnomAD population (Observed) to the number of neutral substitutions expected (Expected) given tri-nucleotide sequence context, CpG methylation levels, and sequencing coverage, using a null model described previously^2^ (**Figure 1**). The Homologous Missense Constraint (HMC) score is defined as the upper bound of the 95% CI of the Observed/Expected ratio. A protein residue with HMC score <1 indicates that missense variants affecting the homologous residues are significantly under negative selection (*P*-value < 0.05) and likely to be deleterious. 3,304,332 possible missense variants (21.7% of assessable) overlie constrained residues with HMC<1. We further classified 1,322,835 possible missense variants (8% of assessable) at highly constrained residues, using a more stringent threshold of HMC<0.8, which we find clinically relevant, as demonstrated below.

**Figure 1.**
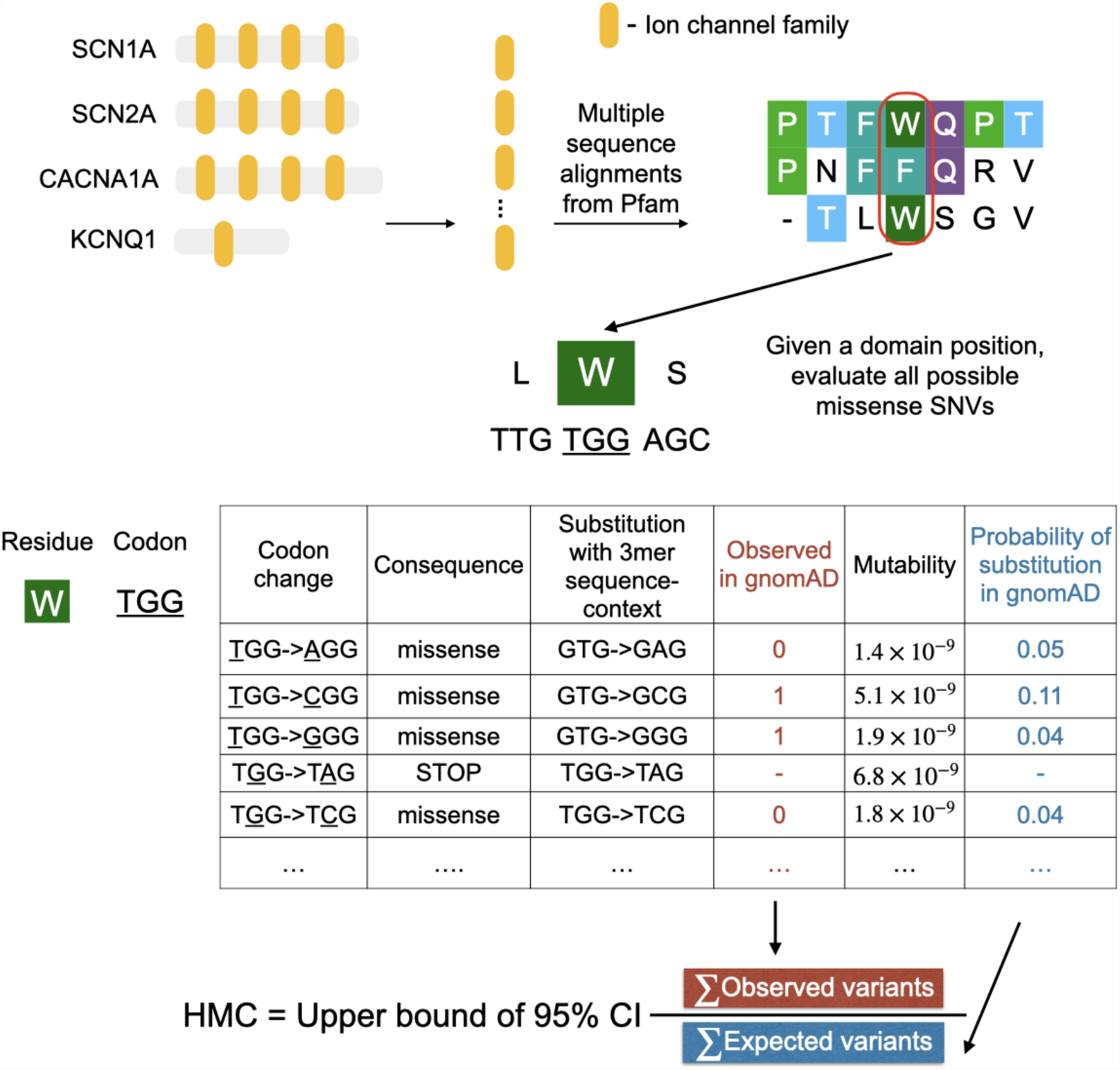
Overview of developing Homologous Missense Constraint. Here we illustrate how to calculate HMC scores using a subset of genes with ion channel domains (Pfam ID: PF00520). Evolutionary equivalent residues were identified by aligning protein sequences across the protein family. Given a domain position with equivalent residues, the Observed/Expected ratio is calculated to measure the genetic constraint of missense variants at this domain position. HMC score is defined as the upper bound of 95% CI of the Observed/Expected ratio. Missense variants affecting domain positions with HMC<1 are significantly (*P-*value < 0.05) under genetic intolerance thus predicted as deleterious.

We robustly evaluated the utility of HMC scores using a wide range of independent tasks, including (i) assessing classification performance of HMC benchmarked against “gold-standard” variant interpretations from ClinVar; (ii) assessing whether HMC prioritises disease-associated variants using case-control analyses in cohorts of patients with known disease phenotypes, without reliance on a gold-standard variant interpretation as a reference; and (iii) evaluating HMC against reference variants evaluated using *in vitro* assays of altered function.

First, we showed that HMC can distinguish pathogenic variants from benign variants in ClinVar. We found that ClinVar pathogenic variants are significantly enriched at constrained domain positions (HMC<1; Rate_Pathogenic vs Benign_=3.9, 95%CI=3.5-4.2, *P*-value=1 ×10^−304^) and significantly depleted at unconstrained domain positions (HMC>=1; Rate_Pathogenic vs Benign_=0.62, 95%CI=0.61-0.64, *P*-value=1 ×10^−311^) (**Figure 2a**). The strength of the association increases as domain residues are under stronger genetic constraint (HMC<0.5; Rate_Pathogenic vs Benign_=37.9, 95%CI=15.7-91.3, *P*-value=1 ×10^−41^) indicating that variants with lower HMC scores are more likely to be disease-causing.

**Figure 2.**
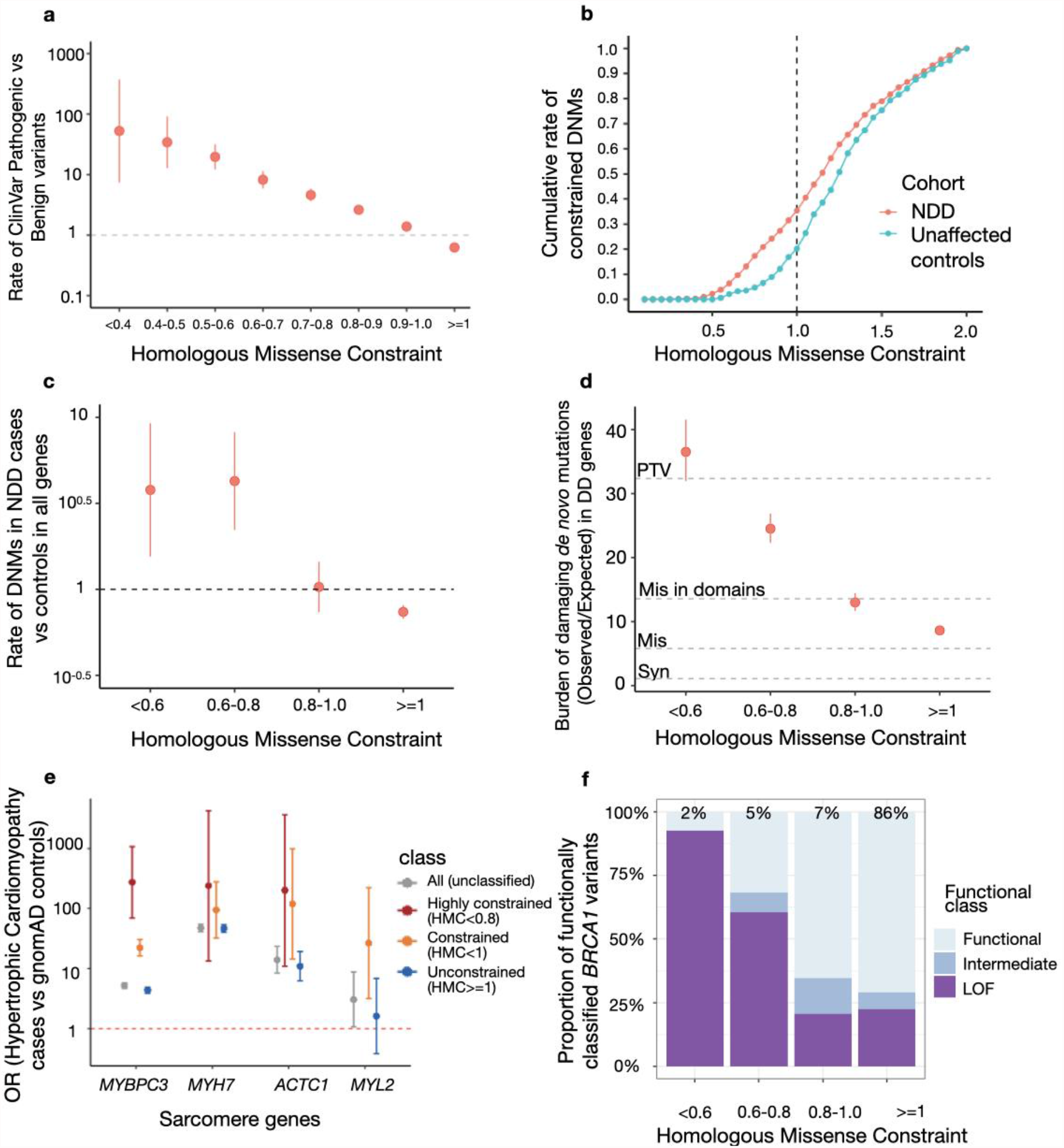
HMC accurately distinguishes pathogenic variants from benign variants. (a) Highly constrained positions within protein domains are enriched for pathogenic variants and unconstrained variants are depleted for pathogenic variants. The rate of ClinVar Pathogenic variants vs Benign variants (Risk ratios) within various HMC constrained/unconstrained bins was plotted with 95% CI. Rate of ClinVar Pathogenic vs Benign variants (Rate_Pathogenic vs Benign_) was calculated as N_in the bin,pathogenic_/N_total,pathogenic_ vs N_in the bin,benign_/N_total,benign_. A total of 13,009 ClinVar Pathogenic and 3,914 ClinVar benign variants (n=3,914) were assessed. (b) Missense *de novo* mutations observed in a cohort of individuals with neurodevelopmental disorders (NDD; n=5,264) are found at more highly-constrained residues than *de novo* mutations observed in unaffected controls (n=2,179). The cumulative rate of constrained *de novo* mutations in cases (N_DNMs with HMC<X in cases_/N_total DNMs in cases_) was plotted to compare with that in controls (N_DNMs with HMC<X in controls_/N_total DNMs in controls_). In total, 1,209 DNMs in cases and 337 in controls are assessed in all genes. (c) Missense *de novo* mutations affecting highly constrained domain positions are significantly enriched in NDD cases versus unaffected controls. The rate of DNMs in cases was compared with that in controls in various HMC constrained/unconstrained bins. Rate of DNMs in cases vs controls was calculated as N_in the bin,cases_/N_total,cases vs Nin the bin,controls_/N_total,controls_. (d) In 285 genes associated with developmental disorders, HMC prioritises damaging *de novo* missense mutations with a comparable effect size as protein-truncating variants (PTV) in 31,058 parent-proband trios of developmental disorders (DD). We compared the prevalence of missense *de novo* mutations (DNM) in established DD-associated genes in individuals with DD against that of expected *de novo* mutations predicted by context-based mutability, and plot the ratio (“burden”) for missense DNMs stratified by HMC score. The burden (Obs/Exp) ratio was calculated as N _observed DNMs in the bin, in cases_/N_expected DNMs in the bin, in cases_. As baseline references, the dotted lines show the burden (Obs/Exp) ratio for synonymous DNMs (OR_Syn_=1.1, 95%CI=1.0-1.2), missense DNMs (without HMC stratification; OR_Mis_=5.8, 95%CI=5.6-6.0), missense DNMs within annotated Pfam domains (OR_Mis in domains_=13.5, 95%CI=12.9-14.3), and protein-truncating DNMs (ORPTV=32.4, 95%CI=30.8-34.0). Missense DNMs at the most highly-constrained residues (HMC<0.6) show an association signal similar to that of protein-truncating DNMs. (e) Highly constrained (HMC<0.8) or nominally constrained missense variants (HMC<1) have increased association with hypertrophic cardiomyopathy compared with controls. We calculated the odds of carrying a rare missense variant for individuals with hypertrophic cardiomyopathy, and for the gnomAD reference population, and show the odds ratio for all rare missense variants, and for rare missense variants stratified by HMC scores.Constrained variants in *MYBPC3* are more strongly disease associated. Data are sparser for the other three genes shown, which are much rarer causes of hypertrophic cardiomyopathy, but the trend is concordant. (f) Highly constrained missense variants are more likely to alter protein function in *BRCA1*. Given *BRCA1* variants classified with *in vitro* assays (LOF: loss-of-function; Functional: functionally neutral; Intermediate: intermediate between the two), we show the proportion of each functional class in HMC constrained/unconstrained bins. Constrained bins include more loss-of-function variants compared with unconstrained bins.

Next, we asked whether HMC could prioritise deleterious *de novo* mutations (DNMs). We analysed published DNMs identified in 5,264 probands ascertained with severe neurodevelopmental delay (NDD) and 2,179 unaffected controls^17^. We found that *de novo* missense mutations in highly constrained domain positions (HMC<0.8) are significantly enriched in NDD cases (Rate_NDD cases vs controls_= 4.1, 95% CI=2.4-6.9, *P*-value=3.1 ×10^−10^; **Figure 2b-c**). Similarly, highly constrained DNMs are significantly enriched in probands ascertained with autism spectrum disorders (Rate_ASD cases vs controls_=2.2, 95% CI=1.3-3.7, *P-*value=0.0028; **Figure S4**). In a larger trio cohort with 31,058 probands of developmental disorders (referred hereafter as “the 31K DD cohort”)^18^, we further evaluated the enrichment of constrained DNMs (the ratio of observed to background expectation^19,20^) in 285 dominant DD-associated genes that showed statistical enrichment of DNMs in that cohort. While missense variants located in annotated domains have a higher burden than those located elsewhere (Obs/Exp=13.6, 95%CI=12.9-14.3), HMC can further narrow down to a subset as highly constrained (<0.8) with an effect size close to that of protein-truncating variants (Obs/Exp=27.6, 95%CI=25.5-29.7 vs PTV: Obs/Exp=32.4, 95%CI=30.1-34.0; **Figure 2d**).

We also tested the ability of HMC to predict deleterious variants causing adult-onset disorders. We performed a case-control gene burden test in 6,327 patients with hypertrophic cardiomyopathy from the SHaRe registry^23^ using the 125,748 gnomAD v2.1.1 exomes as controls. For four sarcomere genes carrying HMC assessable variants, cases have more HMC constrained variants than controls compared to unconstrained or unclassified variants (**Figure 2e**), although due to low variant numbers this is only individually significant for *MYBPC3* (*P*-value=1 ×10^−121^). Genes and variants involved in late-onset phenotypes with smaller effects on reproductive fitness often do not show high signals of genetic constraint. These results suggest that selection signals from early-onset disease genes containing homologous domains could also inform critical variants in late-onset disease genes. Applied to individual genes, we expect HMC would have better statistical power in genes with domains from a large Pfam family with more assessable positions as exemplified here: the I-set (Pfam ID: PF07679) and FN3 (Pfam ID: PF00041) domains in *MYBPC3* belong to large domain families with 785 and 597 homologous copies respectively in the exome while domains from the other three tested genes belong to domain families with no more than 72 copies in the exome (the domain from the largest family: EF-hand_1 (Pfam ID: PF00036) in *MYL2*).

As a further independent evaluation, we compared HMC with functional data from a multiplexed assay of variant effects (MAVE) for *BRCA1*^*21*^. While HMC shows only modest correlation with *BRCA1* MAVE scores in the continuum space (Pearson correlation coefficient *r* = 0.2, reflecting the limited sensitivity of HMC), 70% of highly constrained (HMC<0.8) *BRCA1* variants are loss-of-function, compared to only 23% of unconstrained positions (association between constraint and *in vitro* functional classification OR=8.9, 95%CI=5.4-14.5; **Figure 2f**).

We next compared the performance of HMC against existing pathogenicity scores, using ClinVar variants as a reference set. We first compared HMC to existing sub-genic constraint models: Constrained Coding Region^4^ (CCR), Regional Missense Constraint^5^ (RMC), and a homologous-residue-based conservation metric para_zscore^22^. Using the authors’ recommended thresholds, HMC has better precision than all other methods (**Figure 3a**). We also compared HMC to the state-of-the-art supervised meta-predictors: M-CAP^23^, MPC^5^, REVEL^24^ and CADD^25^. HMC performs comparably to these tools using the highly constrained threshold (<0.8; **Figure 3b**), even though the meta-predictors leverage multiple lines of evidence, and some have been trained directly or indirectly using ClinVar which will tend to inflate their performance, while HMC uses a single line of evidence and is naive to this data.

**Figure 3.**
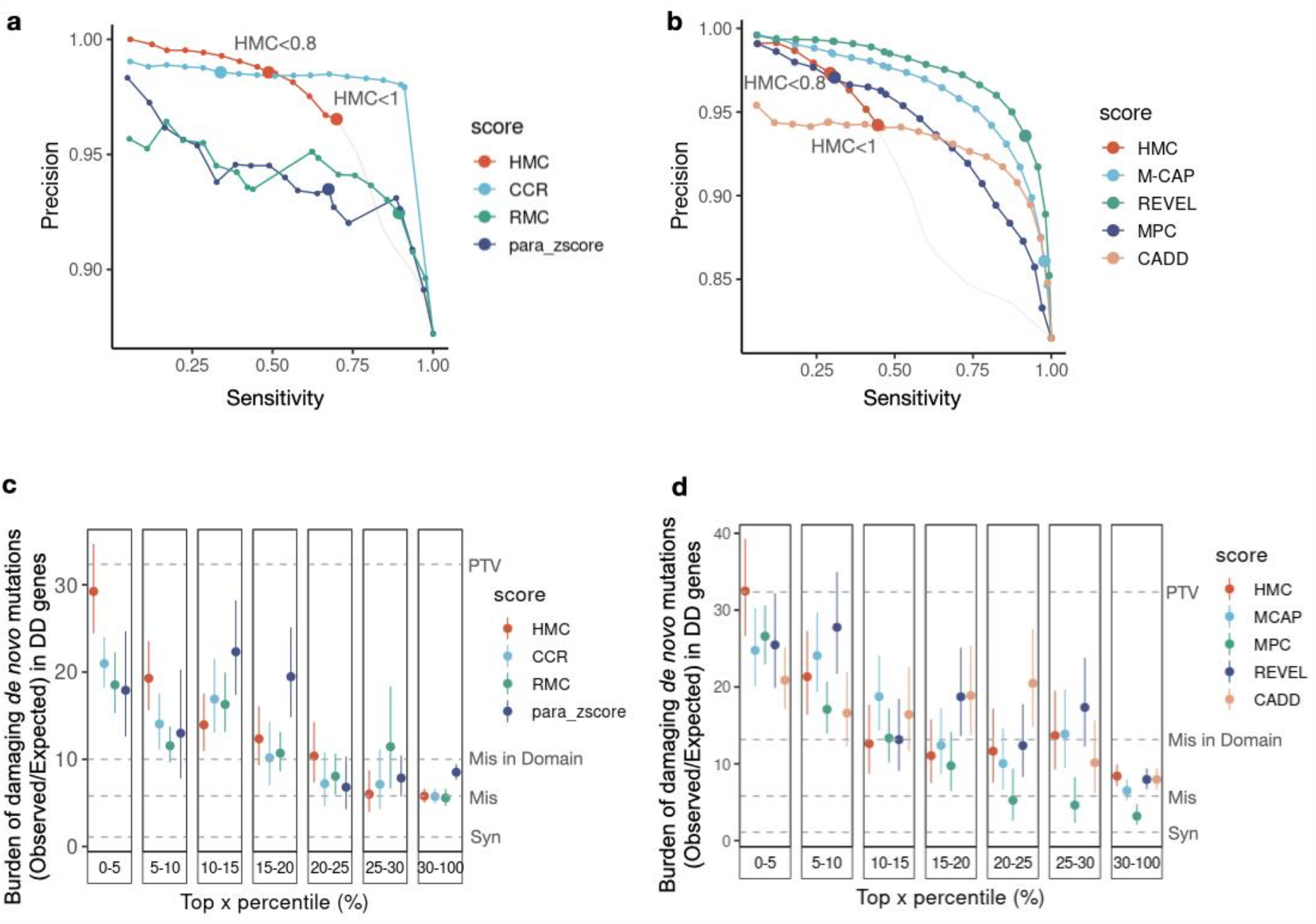
HMC has greater precision than other constraint metrics, and comparable performance to meta-predictor pathogenicity scores. (a) Using ClinVar variants, the Precision-Sensitivity curve demonstrates that HMC has higher precision over the constraint-and homologous-residue-based methods in top-ranked variants and within authors’ recommended thresholds (dots with larger size). The recommended threshold and the corresponding precision and sensitivity for each tool is: HMC<0.8 (98.6%, 48.8%), CCR>95 (98.6%, 34%), RMC<0.6(93.5%, 67.4%) and para_zscore>0 (92.4%, 89.3%). The portion of the curve corresponding to HMC<1 is shown in red, while HMC >1 is shown in pale pink for completeness. As each approach generates predictions on different areas of the exome, we analysed the intersection of ClinVar variants that can be scored by all four methods (3,661 pathogenic and 537 benign variants). (b) HMC has comparable precision as existing state-of-the-art supervised meta-predictors. Dots with larger size indicate the performances (precision, sensitivity) using authors’ recommended thresholds: HMC<0.8 (96.9%, 28.2%), M-CAP>=0.025 (83.6%, 97.2%), REVEL>0.5 (92.2%, 90.8%), CADD >=10(No variants are scored as deleterious), MPC>=2 (97.8%, 28.6%). The performances are reported using 12,428 ClinVar pathogenic variants and 2,824 benign variants, which could be assessed by all the benchmarked methods. (c-d) Using the 31K trio data of DD, the burden of *de novo* mutations (Observed/Expected) in DD-associated genes is compared to evaluate the precision of predicting damaging variants (the higher the burden, the more likely the variants are damaging). For a given tool, variants are grouped into bins based on their percentile of predicted pathogenicity among all assessed variants. (c) Comparison between HMC with existing constraint-based and homologous residue-based scores using DNM burden; (d) Comparison between HMC with existing state-of-the-art meta learners using DNM burden.

We also compared HMC with the above existing pathogenicity scores and constraint models for prioritising deleterious DNMs. Using the 31K DD cohort, HMC outperforms all seven existing tools, being able to identify a subset of DNMs with the highest enrichment in the 285 dominant DD genes (top 5%, **Figures 3c and 3d**), with an effect size as strong as protein-truncating variants. This highlights that HMC is highly effective to distinguish pathogenic variants from bystanders within disease genes.

After confirming HMC is highly precise compared with existing approaches to evaluating missense constraints, we next ask whether it is orthogonal to them. We assessed the distribution of HMC constrained variants across the existing missense constraint metrics including gene-level constraint (MOEUF)^2^, and sub-genic regional constraint scores (CCR and RMC). Constrained homologous residues detected by HMC are distributed across full ranges of these existing metrics in either constrained or unconstrained genes/regions. If a gene/region is more constrained as a whole, on average it also has more constrained residues compared to a less constrained gene/region (**Figure 4**). However, there are substantial numbers of highly constrained missense variants uniquely classified by HMC (<0.8): 893,063 not prioritised by either of the sub-genic metrics, of which 351,175 are not prioritised by any of the existing metrics.

**Figure 4.**
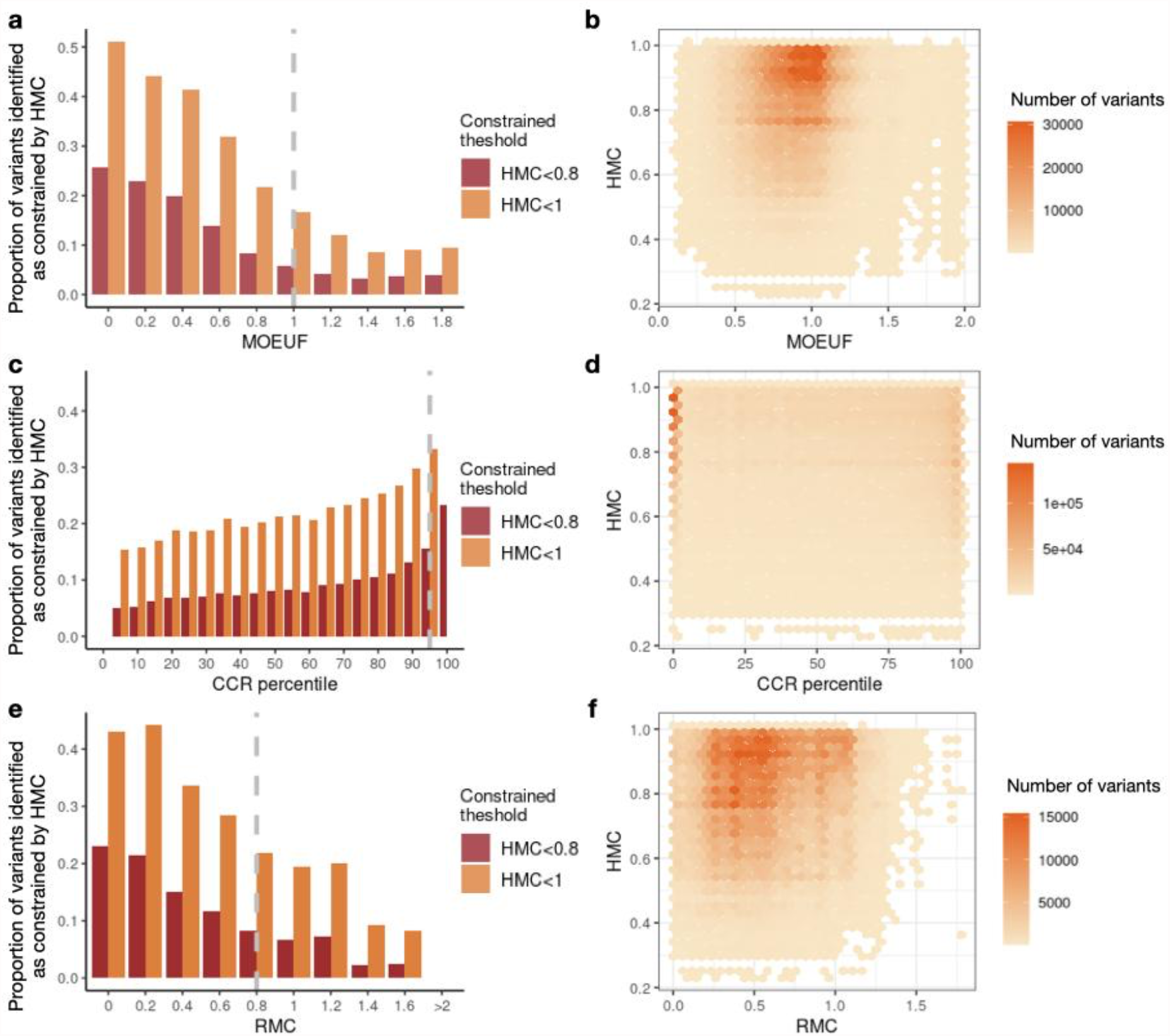
Comparing the distributions of HMC score with existing gene-level and regional level constraint scores. Here we show that HMC is not co-linear with other metrics, and therefore is likely to provide additional information when used in combination. Bar plots in the first column display the proportion of variants identified as constrained by HMC across genes or regions (N_HMC constrained variants in the bin_/N_variants in the bin_). Providing a further detailed view of the bar plots, 2D-histogram plots in the second column display the number of HMC constrained variants within various ranges across gene/region constraint scores. (a-b) The relationship between HMC and a gene’s MOEUF score (gene-level constraint of missense variants; a lower value indicates higher constraint). A gene with MOEUF>=1 (grey dashed line) is considered as nominally unconstrained. (c-d) The relationship between HMC and CCR (a higher percentile indicates higher constraint). A region with CCR percentile <95% (grey dashed line) is considered as unconstrained recommended by authors4. (e-f) The relationship between HMC and RMC (a lower value indicates higher constraint). A region with RMC>0.8 (grey dashed line) is considered unconstrained as recommended by authors.

We further investigate whether HMC could improve gene discovery in developmental disorders given that HMC represents an orthogonal measure of variant deleteriousness and is highly precise to predict disease-causing DNMs in DD genes. HMC prioritises a subset of missense DNMs that show a significant excess burden in the 31K DD probands, in genes that are not previously known to be associated with DD (i.e. not currently considered as diagnostic; HMC<0.8: Obs/Exp=1.37 95%CI=1.25-1.51; **Figure S5**), suggesting its potential to discover unknown DD genes. We updated a gene-specific *de novo* enrichment test (DeNovoWEST)^18^ by incorporating HMC to weight missense variants (see **Methods**) in the 31K DD cohort. Consequently, we observed increased DNM burden of upweighted variants and decreased DNM burden of downweighted ones, indicating the improved separation of pathogenic from benign variants (**Table S1**). Our upgraded tests identified 286 disease-associated genes in the full cohort, and 97 in those previously undiagnosed (probands who do not carry pathogenic variants in consensus diagnostic genes, as previously defined^18^) at genome-wide significance (Bonferroni adjusted *P*-value<0.05/(2 ×18,762)).

Compared with the original study^18^, there are seven newly significant genes across the two tests, which carry at least one constrained missense variant, confirming that their elevated significance signal is driven by HMC (**Table S2-S3**). Four of these genes have previously been published in association with DD via other lines of evidence and are currently included in the Developmental Disorders Genotype-to-Phenotype Database^26^(DDG2P), indicating that our results provide independent support about their gene-disease association. Three of these genes (*BMPR2, KCNC2* and *RAB5C*) have not yet been included in the DDG2P. *BMPR2* is known to cause pulmonary arterial hypertension^27,28^. *KCNC2* has been independently suggested to be a new candidate epilepsy gene^29–31^. Importantly, the newly significant genes all have more constrained missense DNMs than protein-truncating DNMs, suggesting the potential involvement of a gene-function altering mechanism (**Figure 5**).

**Figure 5.**
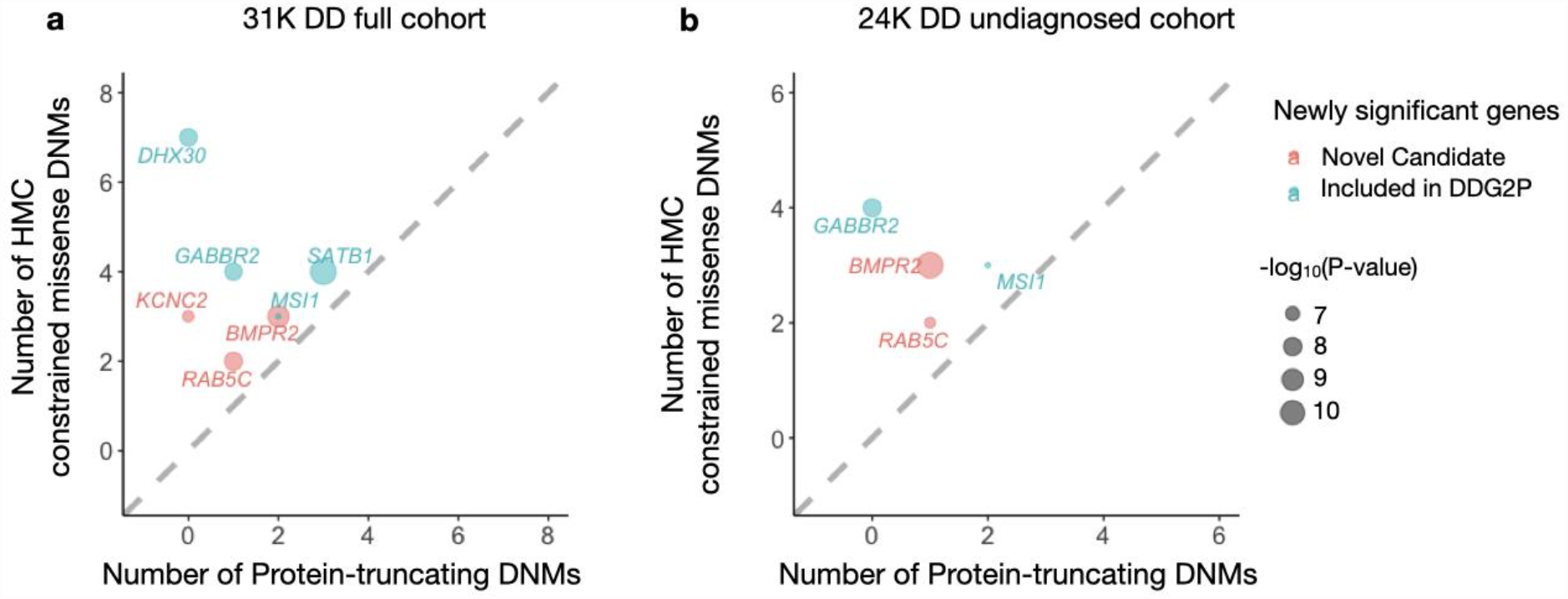
*De novo* variants identified in 31,058 parent-proband trios reveal seven genes associated with developmental disorders at genome-wide significance for the first time in the full DD cohort (a) and the previously-undiagnosed subset (b). Four of these genes have been previously curated as DD genes on the basis of other lines of evidence, and are already included in the G2P database as established Developmental Disorder genes (blue), while three genes represent new candidate DD genes (red). Numbers of constrained missense DNMs classified by HMC and protein-truncating DNMs were compared. The newly-significant associated genes likely act through altered function mechanisms as there are more constrained missense variants than PTVs.

Here we have described a novel measure of genetic intolerance, HMC, to predict deleterious missense variants. Compared with existing metrics that aggregate variants over “horizontal” regions of the genome, HMC considers “vertical” space across homologous regions, enabling us to assess genetic constraint at the single amino-acid level. We demonstrate how HMC is highly precise and orthogonal to existing computational evidence, thus can be broadly used to interpret missense variants and to enhance gene discovery. Though the signal is expected to be driven by genes associated with early-onset disorders under strong negative selection, HMC is informative in adult-onset disease genes, likely leveraging constraint signals from early-onset genes with homologous domains. Although HMC can currently only assess ∼20% of the exome, its statistical power will be increased with the ongoing growth of large-scale population genomics data, efforts of protein family classification, and development of computational structural genomics (providing an alternative definition of homologous variants). HMC provides a powerful new tool for the interpretation of genetic variation.

## Data Availability

All data produced are available online at www.cardiodb.org/hmc.

## Code availability

The essential scripts used to generate HMC scores and recreate the figures in the main text are available at https://github.com/ImperialCardioGenetics/homologous-missense-constraint.

## Data availability

HMC scores for all assessable variants are available via www.cardiodb.org/hmc. External data used in the study were obtained from the following approaches/URLs: The IDs of RefSeq Select Transcripts were downloaded from the UCSC Genome Browser using the Table Browser tool (downloaded date: Nov 28th 2020; options used: group -“Genes and Gene Predictions”, track - “NCBI RefSeq”, table - “RefSeq Select”); ClinVar, https://ftp.ncbi.nlm.nih.gov/pub/clinvar/vcf_GRCh37/archive_2.0/2020/clinvar_20201114.vcf.gz (downloaded at Nov 2020) ; Developmental Disorder Genotype-Phenotype Database (DDG2P), https://www.deciphergenomics.org/ddd/ddgenes (version 2021.11.05); The 31K DD trio data and the original DeNovoWEST, https://github.com/HurlesGroupSanger/DeNovoWEST; gnomAD exome, v2.1.1, https://gnomad.broadinstitute.org/downloads; The hypertrophic cardiomyopathy case series curated by the SHaRe Consortium (data release 2019Q3): https://github.com/ImperialCardioGenetics/CardioBoost_manuscript/tree/master/data/cardiomyopathy/share_variant_count.RData; CCR score, https://github.com/quinlan-lab/ccr; RMC and MPC score, https://storage.googleapis.com/gcp-public-data--gnomad/legacy/exac_browser/regional_missense_constraint.tsv, ftp://ftp.broadinstitute.org/pub/ExAC_release/release1/regional_missense_constraint/fordist_constraint_official_mpc_values.txt.gz; M-CAP score, http://bejerano.stanford.edu/mcap/ (v1.4); REVEL score, https://sites.google.com/site/revelgenomics; CADD, https://cadd.gs.washington.edu/download; para_zscore, https://zenodo.org/record/3582386#.YYxNXb1_rSw (version 9).

## Acknowledgements

We thank Matt Hurles for helpful background discussions. This work was supported by Medical Research Council (UK), British Heart Foundation [RE/18/4/34215], the NIHR Imperial College Biomedical Research Centre, and the Wellcome Trust [107469/Z/15/Z, 200990/A/16/Z]. NW is currently supported by a Sir Henry Dale Fellowship jointly funded by the Wellcome Trust and the Royal Society (Grant Number 220134/Z/20/Z) and funding from the Rosetrees Trust.

For the purpose of open access, the author has applied a CC BY public copyright licence to any Author Accepted Manuscript version arising from this submission. The views expressed in this work are those of the authors and not necessarily those of the funders.

## Methods

### Identification of homologous residues from domain family alignments

The family alignments of all 6,196 human protein domains generated using the NCBI RefSeq sequence database were downloaded from Pfam^15^ (Pfam-A.full.ncbi.gz version 32.0). Given a multiple sequence alignment of a domain family, amino acids in the same column of the alignment were considered homologous.

### Annotation of molecular consequences of variants

RefSeq Select transcripts were used throughout the whole analysis such that each protein-coding gene has a single high-quality representative transcript. The consequences of variants were annotated by VEP (release 101)^32^. Only single-nucleotide variants with VEP annotated as “missense_variant” were included in the analysis.

### Developing a selection-neutral, sequence-context mutational model

To estimate the number of substitutions expected on a single nucleotide, we constructed a neutral mutational model using the gnomAD reference population. Previous studies have shown that the mutation rate of single nucleotide substitution under neutral selection could be predicted based on sequence context and methylation level^1^. Given the baseline substitution rate using a tri-nucleotide sequence text model estimated from variants in intergenic or intronic regions by gnomAD^2^, we calibrated the baseline mutation rate to probabilities of synonymous variants (presumed to be neutral substitutions) within the 125,478 exomes in gnomAD following the procedures described in the gnomAD flagship paper^2^.

We firstly used linear regression to predict the proportions of neutral substitutions given the baseline mutation rates. For each possible tri-nucleotide sequence context, the proportion of neutral substitutions is calculated as the ratio of observed synonymous substitutions over all possible synonymous substitutions. For example, to calculate the proportion of neutral substitutions from AAT to AGT, we firstly find the number of all possible synonymous variants introduced by mutating AAT to AGT along the exome and then count those observed in gnomAD v2 exome data. This ratio of “observed” to “all possible” is used as the dependent variable in linear regression. Since the observation of substitutions would be biased by sequencing coverage, at this step only sites with high coverage (median depth 40) are included in the regression. Two linear regression models were fitted, one for substitutions at CpG sites and the other for non-CpG sites (**Figure S2**). The methylation data for CpG sites was downloaded from gnomAD public datasets and was categorised into three bins: low, medium, and high methylation levels as previously described^2^. With these predicted probabilities of substitutions, we can estimate the expected number of single-nucleotide variants under neutral selection (Expected) in the 125,478 exomes in gnomAD.

Secondly, we adjusted the probabilities of neutral substitutions for low-coverage sites (median depth<40). To this end, the Observed/Expected ratios for synonymous variants were aggregated for each sequencing coverage (measured by median coverage among gnomAD samples). Given a sequencing coverage, it was calculated as: the expected number of variants is the sum of predicted proportions of neutral substitutions for each site derived from the first step, indicating the number we expect with high-coverage sequencing; the observed number of variants is the sum of observed synonymous variants for each site. A linear model was fitted to predict the Observed/Expected ratios given a sequencing coverage on a log10 scale (*R*^*2*^=0.96, *P-value*<2.2×10^−16^; **Figure S3)**. The predicted Observed/Expected ratios by the model were used as correction factors to adjust the expected number of variants at low-coverage sites.

### Estimating Homologous Residue Constraint

An overview of measuring Homologous Residue Constraint is illustrated in **Figure 1**. For an aligned position in a Pfam domain family, we assessed all possible missense substitutions. Among all the possible missense substitutions, the number of substitutions directly observed in gnomAD was counted (Observed). The expected number of missense substitutions was calculated as the sum of predicted probabilities of substitutions given by the neutral mutational model (Expected). The genetic intolerance of this aligned position was calculated as the ratio of Observed/Expected.

Given the variability in the number of aligned domains, in order to control the quality of assessing genetic constraint in homologous residues, we excluded any domain position with less than three expected variants as the number of possible missense variants that occurred at this residue position is too small for us to evaluate genetic constraint robustly. If the number of observed substitutions follows a Poisson distribution under the null hypothesis (no selection), even with zero observed substitutions, the expected number needs to be at least three to reach the significance threshold (the probability of observing zero occurrences with mean occurrence as three is 0.049. In R, it is calculated as “ppois(0,3)=0.049”). It might also indicate the corresponding column is constructed with low confidence filled with a large proportion of gaps (>95% in our observation). Filtering these columns might also limit the effect of alignment bias on defining homologous residues.

Homologous Residue Constraint is defined as the upper limit of 95% confidence interval for the Observed/Expected ratio. The confidence interval for the Observed/Expected ratio was estimated using a Bayesian approach as previously described^2^. The unknown true Observed/Expected ratio (constraint) was considered as a random variable with a uniform prior between 0 and 2 (in computing, discretized into a sequence of 2000 values from 0 to 2, incremented by 0.001). The likelihood function for a given constraint value is given as the Poisson density:

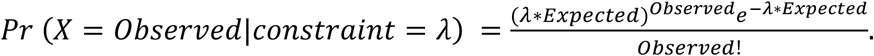

Thus, the posterior probability of a given constraint value could be derived by:

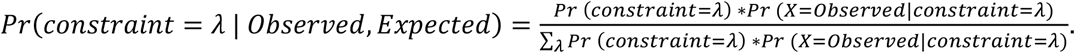

We could further obtain the 95% confidence interval of constraint by taking the 2.5% and 97.5% quantile from its posterior probability distribution. Therefore, the upper bound of 95% CI is taken as the constraint score of homologous residues (HMC). If a residue is scored as HMC <1, it indicates that missense variants disrupting the given domain position are significantly (*P*-value < 0.05) depleted of variants thus under selection pressure.

There are 28,032,394 (∼40% of 70 million possible missense variants) rare (gnomAD MAF<0.1%) missense variants in 15,305 genes (out of 19,212 genes) overlapping 5,807 Pfam domains. After excluding domain positions with limited statistical power, there are 15,236,101 possible rare missense variants from 699 Pfam families with 78,070 domain positions assessable in 9,918 genes. We identified 3,304,332 possible missense variants (21.7% of assessable) at constrained (HMC<1) positions in 596 Pfam domains. 1,322,835 possible missense variants (8% of assessable) were identified at highly constrained residues (HMC<0.8) of 458 domains.

### Evaluating the pathogenicity of ClinVar variants

The association of HMC with known disease-causing variants was tested using ClinVar^33^. The VCF file was downloaded from the ClinVar public FTP site (version 20201114

https://ftp.ncbi.nlm.nih.gov/pub/clinvar/vcf_GRCh37/archive_2.0/2020/clinvar_20201114.vcf.gz). We extracted 22,886 Pathogenic/Likely pathogenic missense variants in Pfam domains, whose clinical significance was recorded as “Pathogenic”, “Likely_pathogenic” or “Pathogenic/Likely_pathogenic”. 7,137 Benign/Likely benign variants in Pfam domains were extracted with clinical significance recorded as “Benign”, “Likely_benign” or “Benign/Likely_benign”. After keeping the HMC assessable domain positions, 13,009 Pathogenic/Likely pathogenic and 3,914 Benign/Likely benign variants were used as test data. Only variants with no conflicting interpretation were included in the test set.

### Evaluating the pathogenicity of *de novo* variants

To test the enrichment of *de novo* variants (DNMs) prioritised by HMC in affected individuals versus unaffected individuals, we analysed the published DNMs in 5,264 patients ascertained with neurodevelopmental disorders, 6,430 patients ascertained with autism spectrum disorder, and 2,179 unaffected controls curated by Satterstrom *et.al*^*17*^.

We applied an independent approach to measure the accuracy of predicting damaging missense *de novo* mutations by testing the enrichment of DNMs prioritised by HMC in affected individuals versus neutral variants estimated by a null sequence-context based *de novo* mutational model^20^. This measurement can also be used to assess whether HMC could distinguish pathogenic and benign variants within disease genes as the enrichment of DNMs in cases vs control individuals can be driven by gene-level disease association. We analysed the published DNMs in 31,058 patients with developmental disorders. The burden of DNMs was calculated as the ratio of the number of observed DNMs to the number of expected DNMs. The number of observed DNMs was directly counted from the variants seen in the cohort. The number of expected DNMs under neutral selection for the cohort is calculated by summing the product of the trinucleotide *de novo* mutation rate and the number of exome samples (2×31,058) for each nucleotide. The *de novo* mutation rate was downloaded from the GitHub repository of denovolyzeR^19^ (https://github.com/jamesware/denovolyzeR-ProbabilityTables/blob/master/data-raw/fordist_1KG_mutation_rate_table.txt). The effective sample size for X-chromosome is adjusted considering sex-chromosome transmission as previously described^18^. Assuming the number of observed DNMs follows a Poisson distribution, the 95% confidence interval for the mean number of observed DNMs could be estimated by using an exact method. In R, it is calculated as “poisson.test(n_obs, conf.level=0.95)”.

To be noticed, the set of DNMs published in Satterstrom *et.al*^*17*^ was a compilation of DNMs from previous publications. Since Satterstrom *et.al*^*17*^ have harmonised the quality control for 5.2K cases and 2.2K unaffected controls thus we used this dataset for DNM case-control analysis. We used the larger set of 31K cases in the DNM enrichment analysis. Of note, the 5.2K cases are a subset of 31K cases^17,18,34^ but we think this won’t affect the validity of our results: the controls are different in the two analyses. 5.2K is compared with unaffected individuals while 31K is compared with DNM null model. Therefore, even if we fully reuse the same cases, these two analyses shall be considered as independent validations.

### Testing improving power of gene discovery

To demonstrate the utility of applying HMC to discover more disease genes reliably, we upgraded the gene-specific *de novo* weighted enrichment simulation test (DeNovoWEST)^18^ by adding HMC to score missense variants. In the original framework of DeNovoWEST, the weight of a missense variant used in the simulation test depends on the regional missense constraint (RMC). Here we incorporated HMC into this framework through the following procedures: (1) combining HMC with regional missense constraint to label constrained missense variants, thus a missense variant is considered as constrained if either RMC or HMC (HMC<0.8) score it as constrained. Compared with the original version, there are 732,404 more missense variants classified as constrained; (2) updating the weights of missense variants used in DeNovoWEST: we calculated the burden of *de novo* missense variants against a null *de novo* mutational model^20^ and inferred the corresponding positive predictive values (PPV) for all possible categories using constraint (based on step 1) and CADD scores. The newly derived PPV is used as weights in the downstream gene-specific test. The upgraded test was applied separately in the full (n=31,058) and undiagnosed (n=24,288) cohort of parent-proband trios of developmental disorders^18^. Compared with the original version, the DNM burden of constrained missense variants have increased in the 31K DD cohort while the one of unconstrained missense variants have decreased (**Table S1**). This suggests that HMC has improved the classification of pathogenic and benign missense DNMs in the cohort.

We define newly-significant associated genes driven by HMC scoring that meet all the following criteria: (1) only reached genome-wide significance threshold (Bonferroni adjusted *P*-value<0.05/ (2 ×18,762)) in our upgraded test; (2) carries at least one highly constrained missense variant prioritized by HMC (HMC<0.8). In our upgraded tests, there are seven genes in total that reached genome-wide significance level but did not in the original DeNovoWEST tests: *BMPR2, DHX30, GABBR2, KCNC2, MSI1, RAB5C, SATB1* (**Table S2-S3**). All of them have at least one HMC constrained missense variant thus are defined as newly-significant associated genes in our analysis. There are three genes (*ARHGEF9, SETD1B* and *CNKSR2)* that were significant in the original tests but did not in the upgraded tests. Since the significance levels of these three genes before and after the upgrade are in the same scale, we think it’s likely due to random variations in different runs of simulations. For those currently not included in DDG2P, we define them as novel candidate genes associated with DD including *BMPR2, KCNC2* and *RAB5C*.

### Compare HMC with functional assay data

We collected mutagenesis experiment data on missense SNVs of 13 exons of *BRCA1* RING domain^21^. Variants are functionally classified as: functionally-neutral, intermediate, loss-of-function. We were able to analyse 1321 out of 3893 missense SNVs in the assay data.

## Notes

### Competing Interest Statement

The authors have declared no competing interest.

### Author Declarations

This study involves only openly available human data, which can be obtained from: gnomAD, ClinVar, code repository (https://github.com/ImperialCardioGenetics/homologous-missense-constraint) and references indicated in the manuscript.

