## Supplementary Material for "Genetic constraint at single amino acid resolution improves missense variant prioritisation and gene discovery"

### Supplemental Contents

**Supplementary Figures** 2

**Supplementary Tables** 8

**Supplementary Information** 11

#### Supplementary Figures

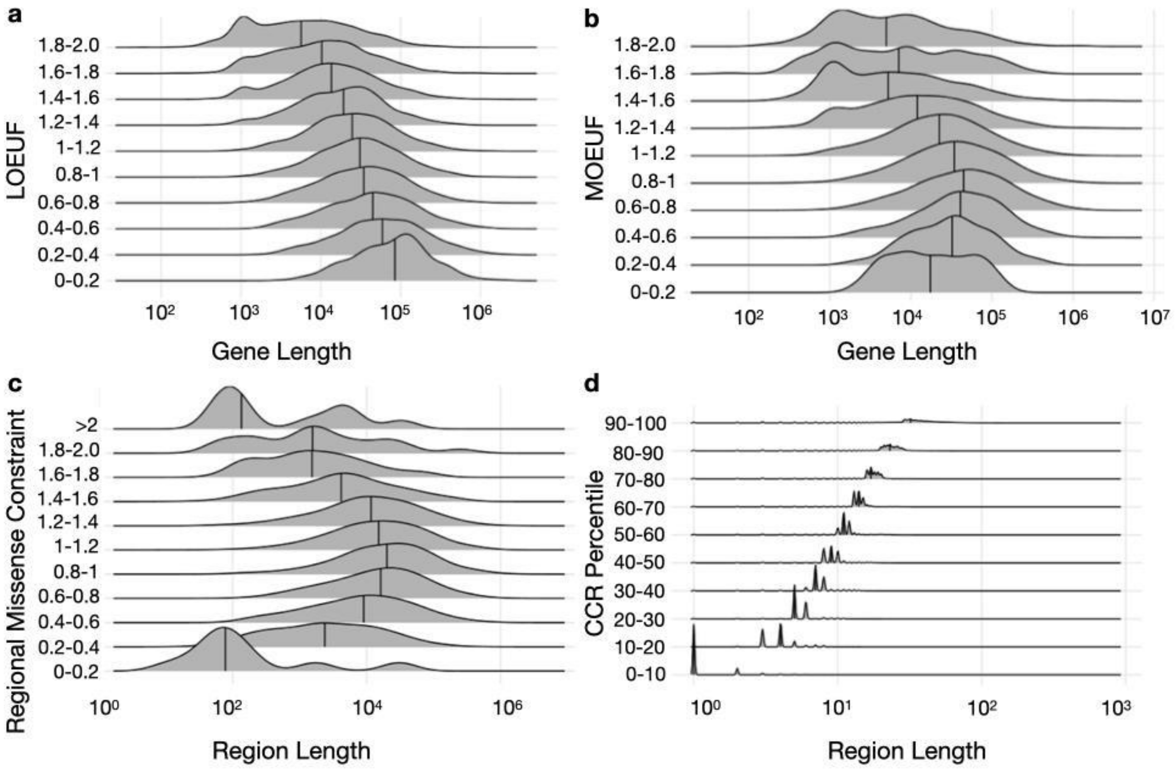


**Figure S1. The relationship between the length of a coding region and its genic or regional level constraint scores.** We demonstrated the relationship using four existing genic and regional constraint scores: LOEUF[1] (a), MOEUF[1] (b), Regional Missense Constraint (RMC)[2] (c) and CCR[3] (d). The probability density of the length of a coding region and the median (indicated as the vertical line) across different ranges of constraint scores are shown. Constrained bins could be loosely defined as: LOEUF<1, MOEUF<1, RMC<1, CCR Percentile>90. Since these existing metrics measure the constraint signals linearly along the exome, in most cases, their constrained bins include longer coding sequences likely clustered with pathogenic variants than the unconstrained bins. Regions/Genes with short sequences or pathogenic variants sparsely distributed could be missed out as unconstrained.

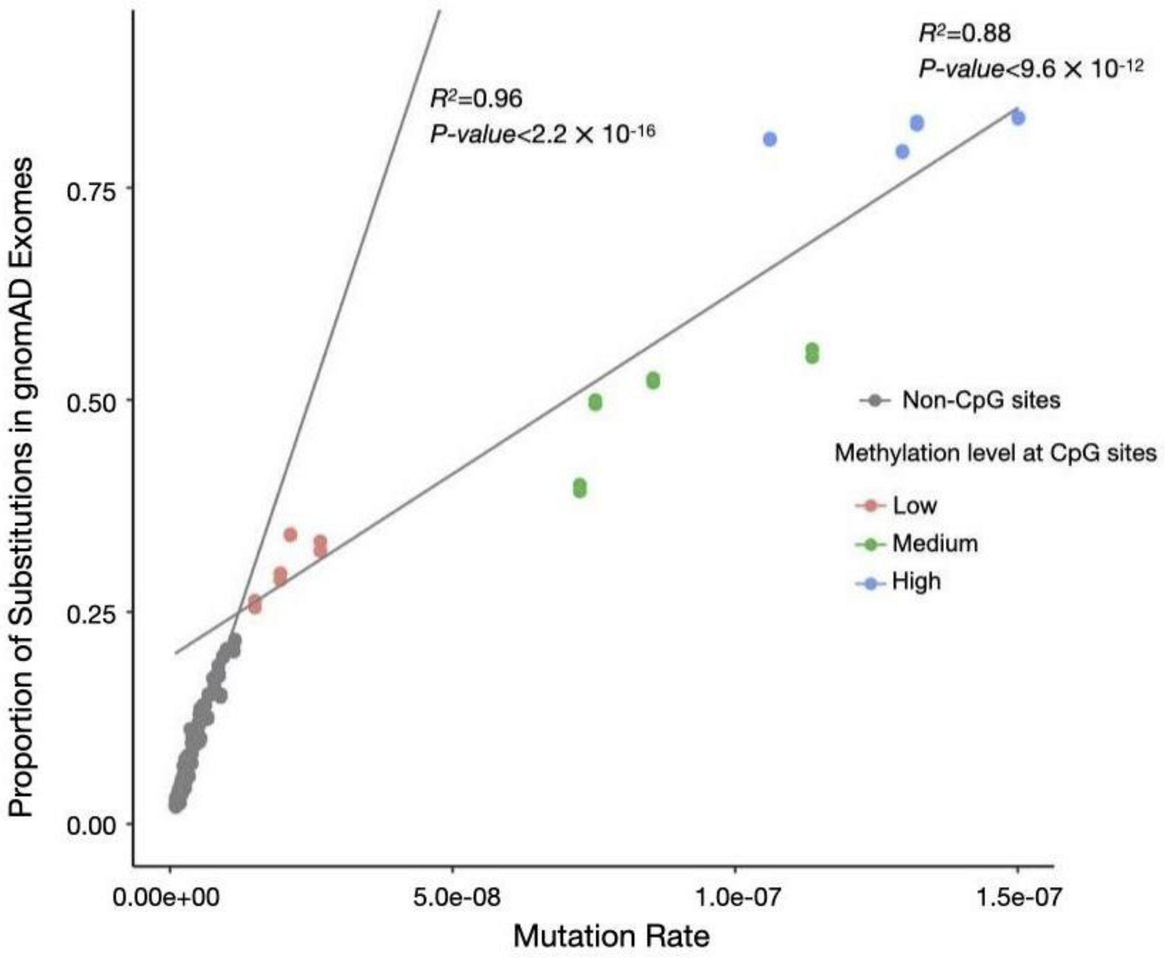


**Figure S2. Calibration of baseline mutation rates to probabilities of neutral substitutions.** Two linear regression models were fitted to predict the proportions of neutral substitutions within the 125,478 exomes from gnomAD: one for CpG sites and the other one for non-CpG sites. This shows that the model is well calibrated for the effect of CpG methylation. In the plot, each dot represents a type of substitution specified by trinucleotide sequence context and methylation level (for CpG sites).

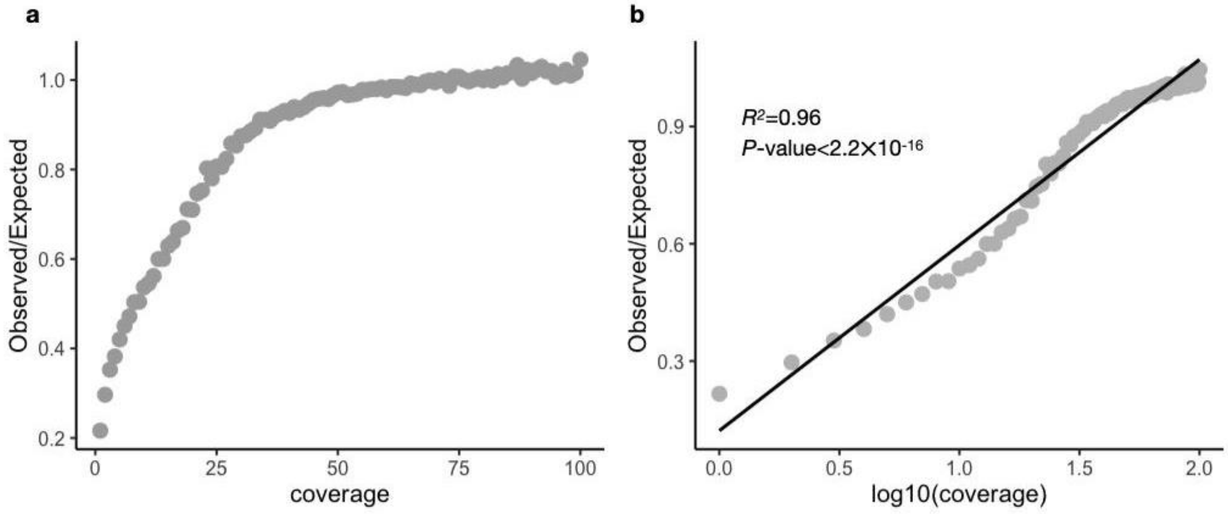


**Figure S3. Calibration of probabilities of neutral substitutions on low-coverage sites (coverage<40).** (a) The relationship between sequencing coverage and Observed/Expected ratios. (b) A linear model is fitted to predict the Observed/Expected ratios given a sequencing coverage on $\log_{10}$ scale. The predicted Observed/Expected ratios were used as correction factors to adjust the expected number of variants at low-coverage sites.

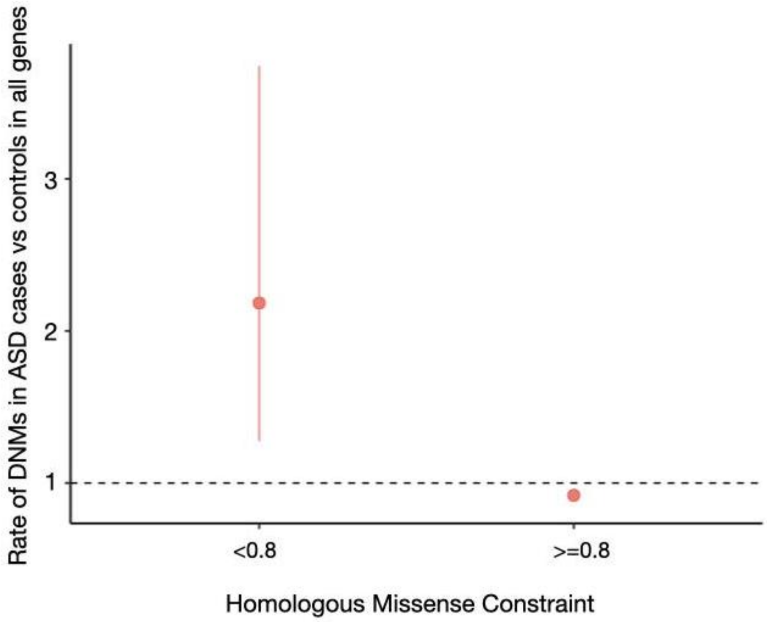


**Figure S4. Enrichment of constrained missense DNMs in 6,430 patients ascertained with autism spectrum disorders versus 2,179 unaffected controls.**

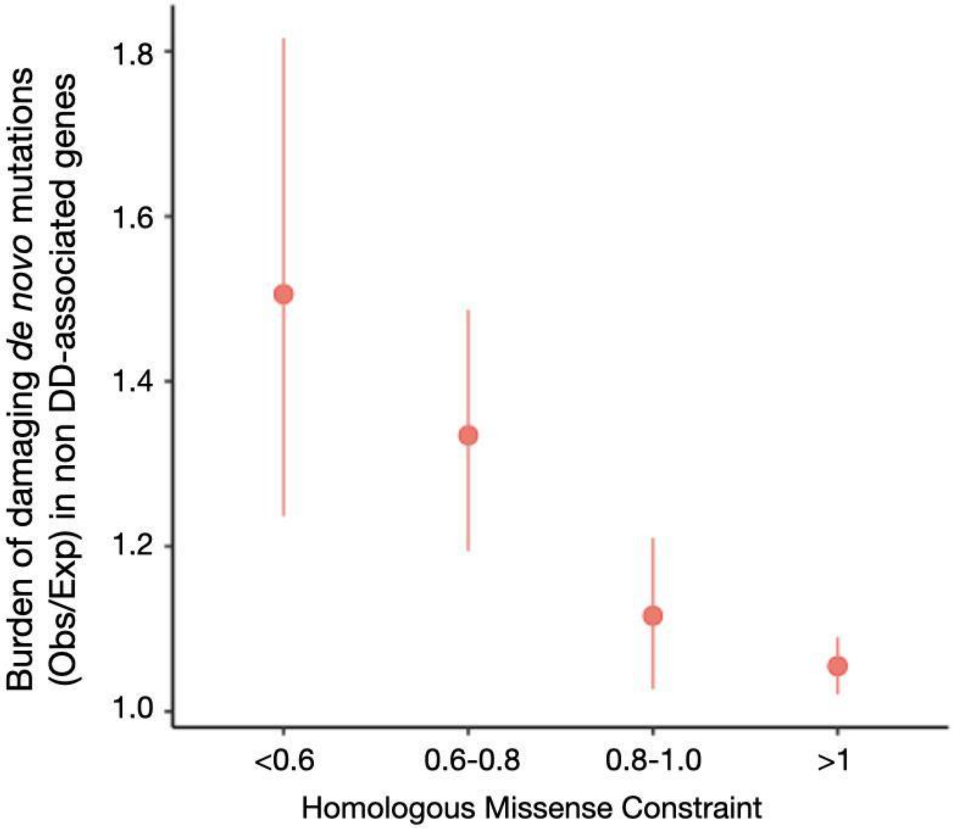


**Figure S5. Enrichment of constrained missense DNMs of non DD-associated genes in 31K DD trios.** 18,644 genes not considered in diagnostics are defined as non-DD genes as described in the publication of 31K DD cohort[4].

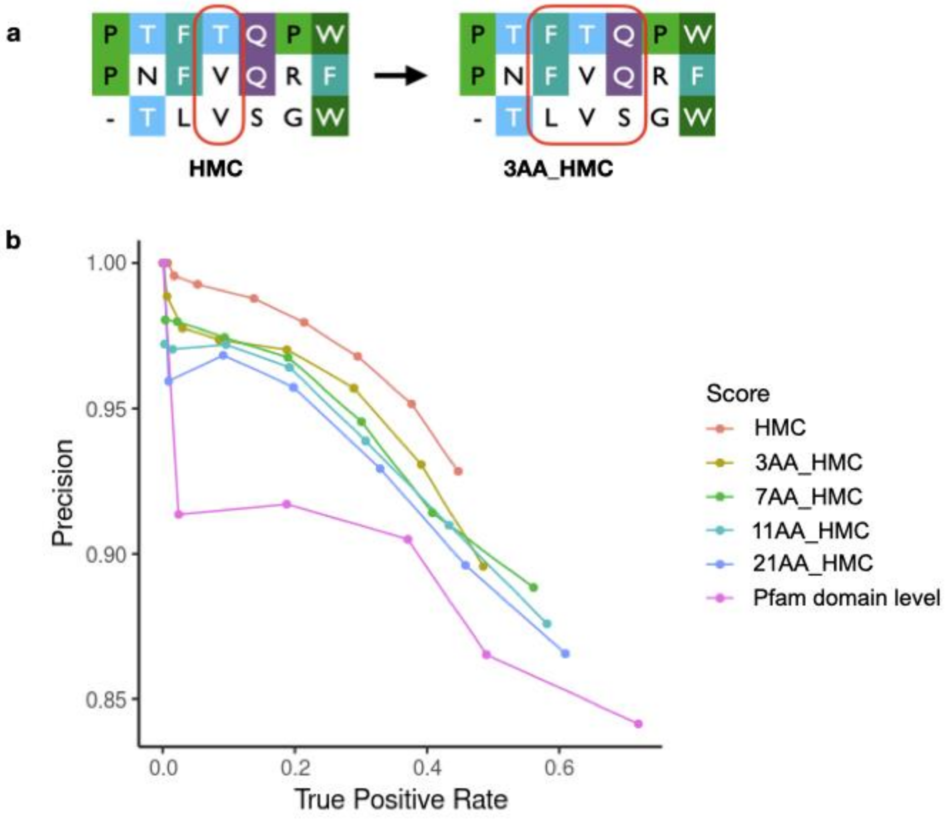


**Figure S6. Exploring alternative genetic constraints measured in Pfam domains.** The following metrics are calculated: HMC (vanilla), 3AA_HMC (Constraint of homologous residues within a sliding window of 3 amino acids, illustrated in **a**), 7AA_HMC, 11AA_HMC, 21AA_HMC and genetic constraint of domain-level. Their performance was compared by classifying ClinVar interpreted data shown as Precision-Recall curves in **b**. HMC has the best precision over all the alternative methods when the constraint scores are under 1.

#### Supplementary Tables

**Table S1. Comparison of DNM burdens of constrained and unconstrained missense variants in the upgraded and original DeNovoWEST tests in the 31K DD cohort.** In DeNovoWEST, missense variants are subsetted based on missense constrained information, CADD scores and $S_{het}$ values ("estimated selective effect of heterozygous PTVs on gene level") and scored according to observed DNM burdens (obs/exp ratio) in the 31K DD cohort. Compared with original version of DeNovoWEST, after we incorporated HMC to score missense variants, constrained missense variants have increased enrichments while unconstrained missense variants have decreased enrichments, indicating that HMC improved the discrimination of missense variants associated with diseases.

| | DNM burden and 95%CI after upgrading | DNM burden and 95%CI in the original publication[4] |
|---|---|---|
| Constrained missense variants | 2.661 [2.581-2.743] | 2.597 [2.513-2.684] |
| Unconstrained missense variants | 1.127 [1.113-1.141] | 1.145 [1.130-1.159] |

**Table S2. Newly-significant DD-associated genes found in the full cohort of 31K DD trios.**

| Gene | *P*-value in original DeNovoWEST | *P*-value in updated DeNovoWEST | Number of PTVs | Number of missense variants (constrained/total) | DDG2P Confidence in Monoallelic mode* (version 2021.12.12) |
|---|---|---|---|---|---|
| *BMPR2* | 6.25e-06 | 9.59e-10 | 2 | 3/4 | No |
| *DHX30* | 2.62e-06 | 1.63e-08 | 0 | 7/8 | Probable |
| *GABBR2* | 3.56e-06 | 1.78e-08 | 1 | 4/8 | Probable |
| *KCNC2* | 3.12e-03 | 2.64e-07 | 0 | 3/4 | No |
| *MSI1* | 2.74e-05 | 6.53e-07 | 2 | 3/5 | Possible |
| *RAB5C* | 2.12e-04 | 1.13e-08 | 1 | 2/5 | No |
| *SATB1* | 2.22e-06 | 1.65e-11 | 3 | 4/7 | Confirmed |

* - The terminology used to describe gene-disease validity is explained here

https://www.ebi.ac.uk/gene2phenotype/terminology

**Table S3. Newly-significant DD-associated genes found in the undiagnosed cohort of 24K DD trios.**

| Gene | *P*-value in original DeNovoWEST | *P*-value in updated DeNovoWEST | Number of PTVs | Number of missense variants (constrained/total) | Status in DDG2P in Monoallelic mode (version 2021.12.12) |
|---|---|---|---|---|---|
| *BMPR2* | 2.44e-06 | 1.26e-10 | 1 | 3/4 | No |
| *GABBR2* | 3.56e-06 | 8.34e-09 | 0 | 4/7 | Probable |
| *MSI1* | 6.47e-06 | 1.16e-07 | 2 | 3/5 | Possible |
| *RAB5C* | 1.19e-03 | 7.07e-08 | 1 | 2/4 | No |

#### Supplementary Information

##### Comparison with existing works

The idea of aggregating genetic intolerance information over homologous residues in protein domains has been developed previously by Wiel and colleagues[5,6], who measured the genetic intolerance of homologous positions as non-synonymous over synonymous ratio (or $d_N/d_S$). Here, we have adopted an improved measure of genetic intolerance, which uses a sequence context-based mutability model as a neutral control (genetic constraint throughout the manuscript) instead of relying on empirical observation of synonymous variants. Genetic constraint has been shown extensively to improve the statistical power compared with $d_N/d_S$ ratio or other genetic intolerance measures[7–9].

##### Discussion on alternative approaches to develop HMC

Here we discussed several ways one could develop HMC alternatively and the consequences.

Since there is no gold-standard definition for homologous amino acids, our choice is largely limited by the availability of data. In this study, we used protein domain alignment to define homologous residues because there are more genes annotated with protein domains compared with paralogous alignment and structural alignment. For 19,212 genes included in RefSeq Select, 15,305 have Pfam domains while 14,772 genes have paralogs. For structurally-aligned residues, currently there are not yet standard resources publicly available. Given that only ~50% of human proteins have structural models including experimentally determined structures and homology-based predicted structures[10], the number of genes that could be reliably structurally aligned with others would be even less. However, one could revisit this comparison when the size of data goes up in the future.

As the performance of HMC could be affected by the bias of sequence alignment, we also explored whether taking account of the genetic constraint of surrounding amino acids could improve the performance since the true homologous residues are likely in neighboring columns if not aligned with each other. Our experiment shows that adding more surrounding amino acids could improve sensitivity but also compromise precision (positive predictive value) since there could be more non-relevant residues simultaneously added to dilute the signal (**Figure S6**). To favor precision over sensitivity, we did not consider adding surrounding amino acids in our final metric and used the vanilla version.

Additional features of residues could be added to improve the positive predictive value of HMC, such as interspecies conservation and biochemical properties of aligned amino acids. As homologous residues based on sequence might not be always functionally homologous to each other, the performance of HMC could be also affected by exceptions when certain individual residues might have different functional consequences/specifications than homologous residues in their family. Though we chose to keep HRC orthogonal here without adding existing molecular evidence, there is potential for improvement by combining HMC with additional features of residues.
